# Data Visualization for Surgical Informed Consent to Communicate Personalized Risks and Patient Preferences

**DOI:** 10.1101/2020.03.25.20038398

**Authors:** Undina Gisladottir, Drashko Nakikj, Rashi Jhunjhunwala, Gabriel Brat, Nils Gehlenborg

## Abstract

**Objective:** Identify key elements of an effective visualization method for communicating personalized surgical risks to patients.

**Background:** Currently, there is no consensus on which risks should be communicated during the informed consent process and how. Furthermore, patient preferences are often not considered during the consent process. These inefficiencies can lead to non-beneficial outcomes and raise the potential for legal implications. To address the limitations of the informed consent process, we propose a visual consent tool (VCT) that incorporates patient preferences and communicates personalized risks to patients using data visualization.

**Methods:** To understand how patients perceive risk visualizations and their role in the informed consent discussion, we gathered feedback on visualizations by conducting semi-structured interviews during postoperative visits. Thematic analysis was performed to identify major themes. Iterative evaluation and consolidation of the major themes were performed with domain experts.

**Results:** A total of 20 patients were interviewed for this study with a median age of 59 (sd = 14). The thematic analysis revealed factors that influence the perception of risk, of risk visualizations, and the usefulness of the proposed VCT. We found that patients preferred VCT over the current methods and had different preferences for risk visualization. Further, our findings suggest that surgical concerns of patients were not in line with existing risk calculators.

**Conclusion:** We were able to identify key elements that influence effective risk communication in the perioperative setting. We found that patient preference is variable and should influence choices for risk presentation and visualization.

## INTRODUCTION

In the United States more than 50 million surgical procedures are performed annually (“NQF: Surgery 2015-2017 Final Report” n.d.). For each procedure, a clinician acquires informed consent from the patient or a surrogate. The discussion during this process plays an important legal and ethical role and should determine the appropriate treatment plan for each patient. Literature suggests that this discussion often does not address the patient’s personal treatment goals. Additionally, many important details are solely communicated verbally. Inadequate unexpected, and possibly life threatening, events can lead to malpractice lawsuits (Cooper et al. 2014; Bickmore et al. 2015; Grauberger et al. 2017). Instead, the informed consent conversation should properly set the patient’s expectations to decrease the chances of what a patient would consider a non-beneficial outcome (Cooper et al. 2014; Angelos 2012; Grauberger et al. 2017).

Although medical professionals agree that determining patient priorities is important for choosing the appropriate treatment plan, the discussion during the informed consent often fails to consider the patient’s condition and treatment goals (Cooper et al. 2014; Angelos 2012). Furthermore, the current informed consent process is not standardized and leaves patients without a clear understanding of the consequences of surgery (Scheer et al. 2012). There is also a lack of consensus in the medical community of which risks to communicate and risk estimates are often too broad and vary between physicians (Cooper et al. 2014). Multiple studies have shown that despite reviewing the surgical procedure and associated risks, the patients’ understanding after these discussions is well below acceptable limits (Fink et al. 2010). Risk score calculators try to expand the conversation through personalized risks for any given patient. They provide discrete risk scores for a variety of outcomes based on the surgical procedure and preoperative patient data. Despite the growing prevalence of these tools, the surgical community has not reached a consensus on how to communicate these scores. Some groups have attempted to address this issue by categorizing the complications into Best Case/Worst Case or Good/Intermediate/Bad (Kruser et al. 2015; Taylor et al. 2017; Ingraham et al. 2017). For these approaches, patient preference, which is essential to defining a good outcome for a patient, is not necessarily incorporated or used to inform the conversation.

We propose a design for the Visual Consent Tool (VCT) to address previous limitations and (1) incorporate patient preferences, (2) set expectations for the upcoming surgery, and (3) standardize risk communication during informed consent. The VCT communicates personalized risks to patients in three main steps (Figure 1). First, personalized risks are calculated using one of the existing risk prediction models currently available (Bertsimas et al. 2018; Meguid et al. 2016; Bilimoria et al. 2013; Brennan et al. 2019). These prediction models typically incorporate a surgical CPT code and patient preoperative data to calculate risks. Second, patients select a limited number of major concerns (we chose 3 arbitrarily) out of a list of twenty complications pre-ranked in descending order of likelihood. Finally, we visualize the probability of the three most likely and patient-selected complications as well as the potential discharge destinations: home, rehabilitation, and death. With this, the VCT allows patients to compare the risks of most likely and prioritized complications and communicates potential discharge destinations after surgery.

**Figure 1:**
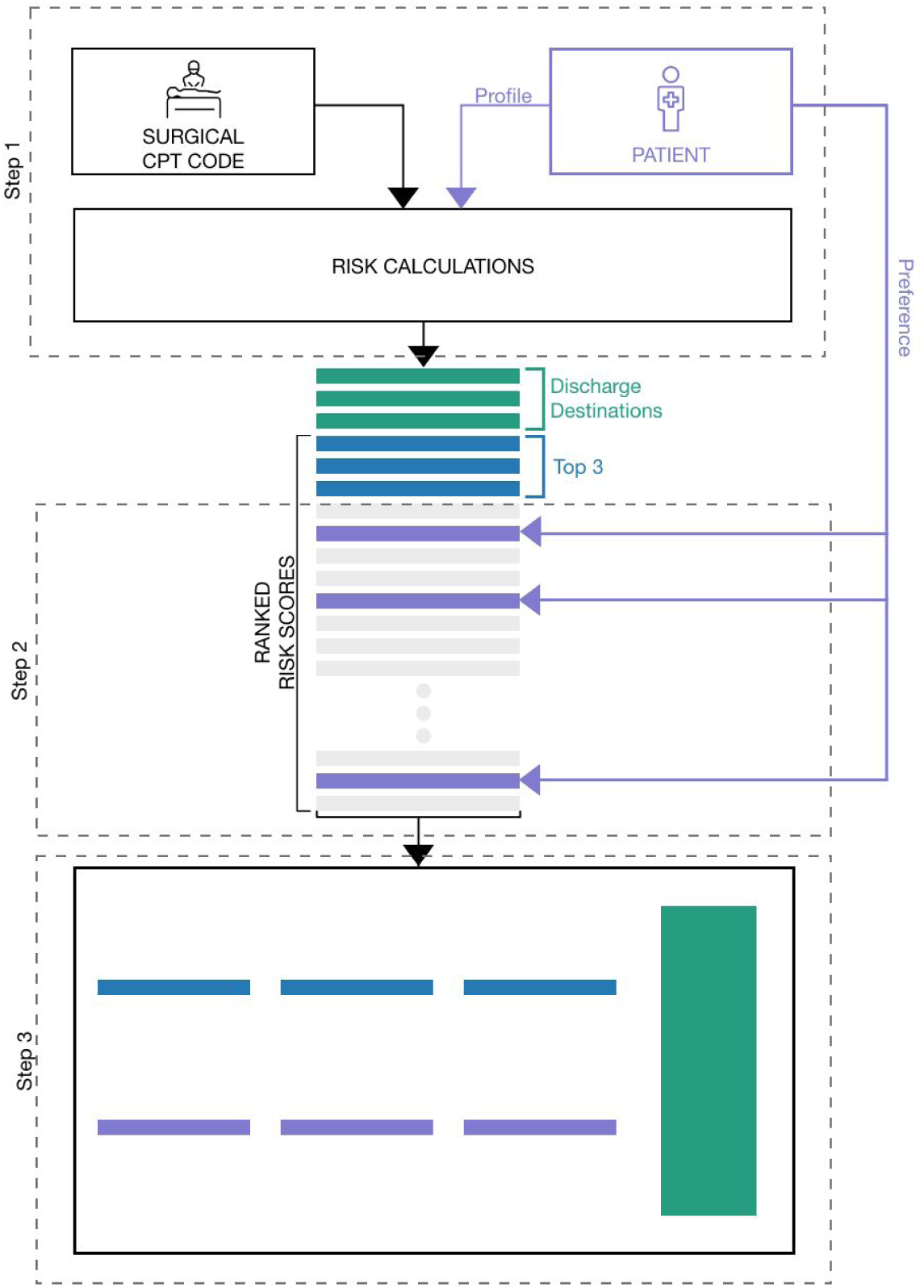
The proposed Visual Consent Tool (VCT) includes three main steps to help the patient and surgeon determine whether surgery is the appropriate option or other alternatives should be explored. In the first step, personalized risks are calculated using an existing risk model with identification of surgery (e.g. CPT code) and inputs of patient preoperative data. The patient then chooses up to three risks that are of highest concern (purple bars) in addition to the top three (blue bars). Finally, a visualization of these six risks is displayed along with the likelihood of each of the final discharge destinations (green bar).

Whereas multiple models for personalized risk calculation exist, the major focus of this study was to understand how to communicate these personalized risks using data visualization (Bilimoria et al. 2013; Bertsimas et al. 2018; Meguid et al. 2016; Brennan et al. 2019). To assess how well the VCT is able to set expectations and whether it can improve the informed consent discussion, we conducted a mock-up based evaluation of the VCT’s personalized risk visualization component. We conducted semi-structured interviews with patients during their postoperative visit to the acute care surgical clinic at an academic medical center. Through thematic analysis of the interviews, we identified several factors that impact the perception of risk, perceived value of risk visualization, the effects of the risk visualization and the potential usefulness of the VCT in a real life setting. The report of this study is based on the consolidated criteria for reporting qualitative research (COREQ) (Tong, Sainsbury, and Craig 2007).

## METHODS

### Visual Consent Tool (VCT) Design

A schematic overview of the Visual Consent Tool (VCT) is shown in Figure 1. The VCT consists of 3 elements: risk calculation, preference identification, and risk visualization.

#### 1. Risk Calculation

Multiple methods of calculating personalized perioperative risks for patients have been published (Bilimoria et al. 2013; Bertsimas et al. 2018; Meguid et al. 2016; Brennan et al. 2019). These calculators use collected patient data (e.g. age, gender, and smoking status) to calculate risk of a given post-operative complication. As an example, the American College of Surgeons (ACS) risk calculator, the most commonly used tool, leverages National Surgical Quality Improvement Program (NSQIP) participant data over 400 hospitals to calculate 20 different perioperative risks (Bilimoria et al. 2013). In this study, we did not focus on improving risk calculation. Generally, our approach could be applied to risks calculated with any risk calculator. For practical purposes, we used results obtained from the ACS risk calculator for this study. In our proposed interface, the surgeon provides information about the surgery by entering a CPT code and the patient profile (Figure 2).

**Figure 2:**
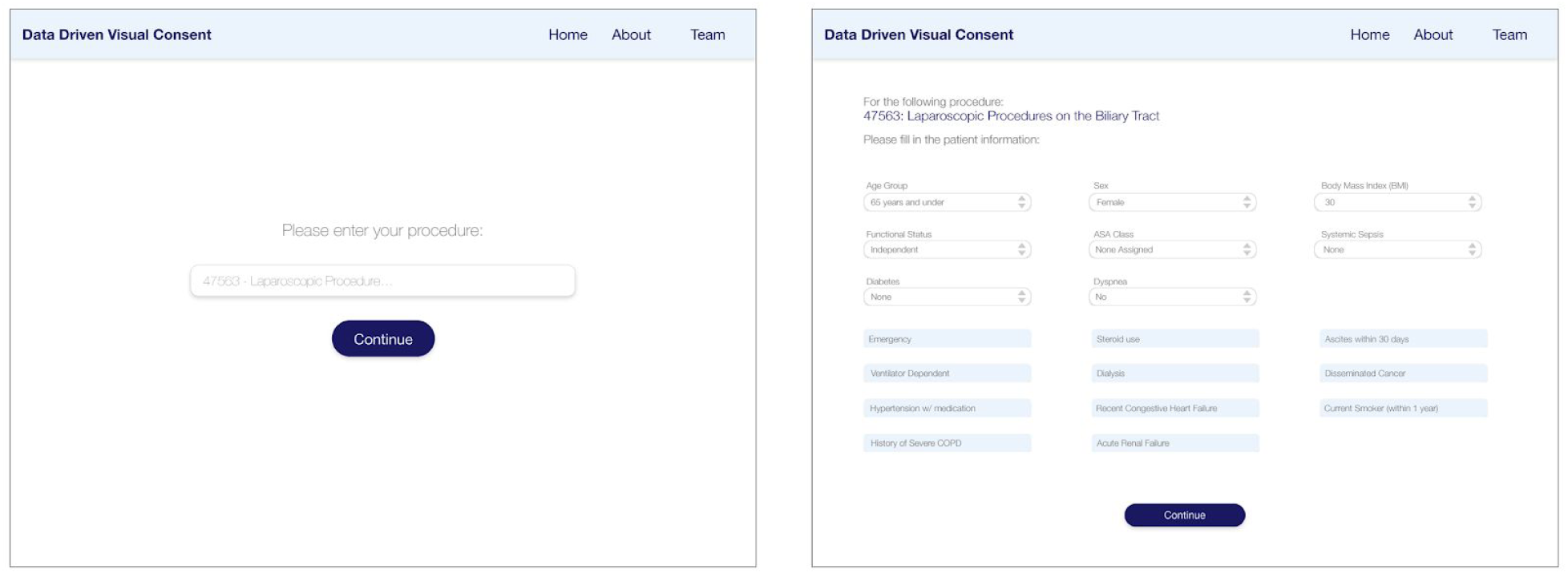
Patient Profile and CPT code input. The form on the left is used to enter the CPT code for the surgery and the form on the right is used to provide patient characteristics required by the risk calculator.

#### 2. Incorporating Patient Preference

To incorporate personal preferences, our tool provides an interface for patients to identify three complications that are of particular concern, in addition to the top three risks that the tool automatically selects based on the risk calculations (Figure 3). After presenting the patient with a list of possible complications pre-ranked by likelihood, patients are able to choose the risks that are most important to them (Figure 3).

**Figure 3:**
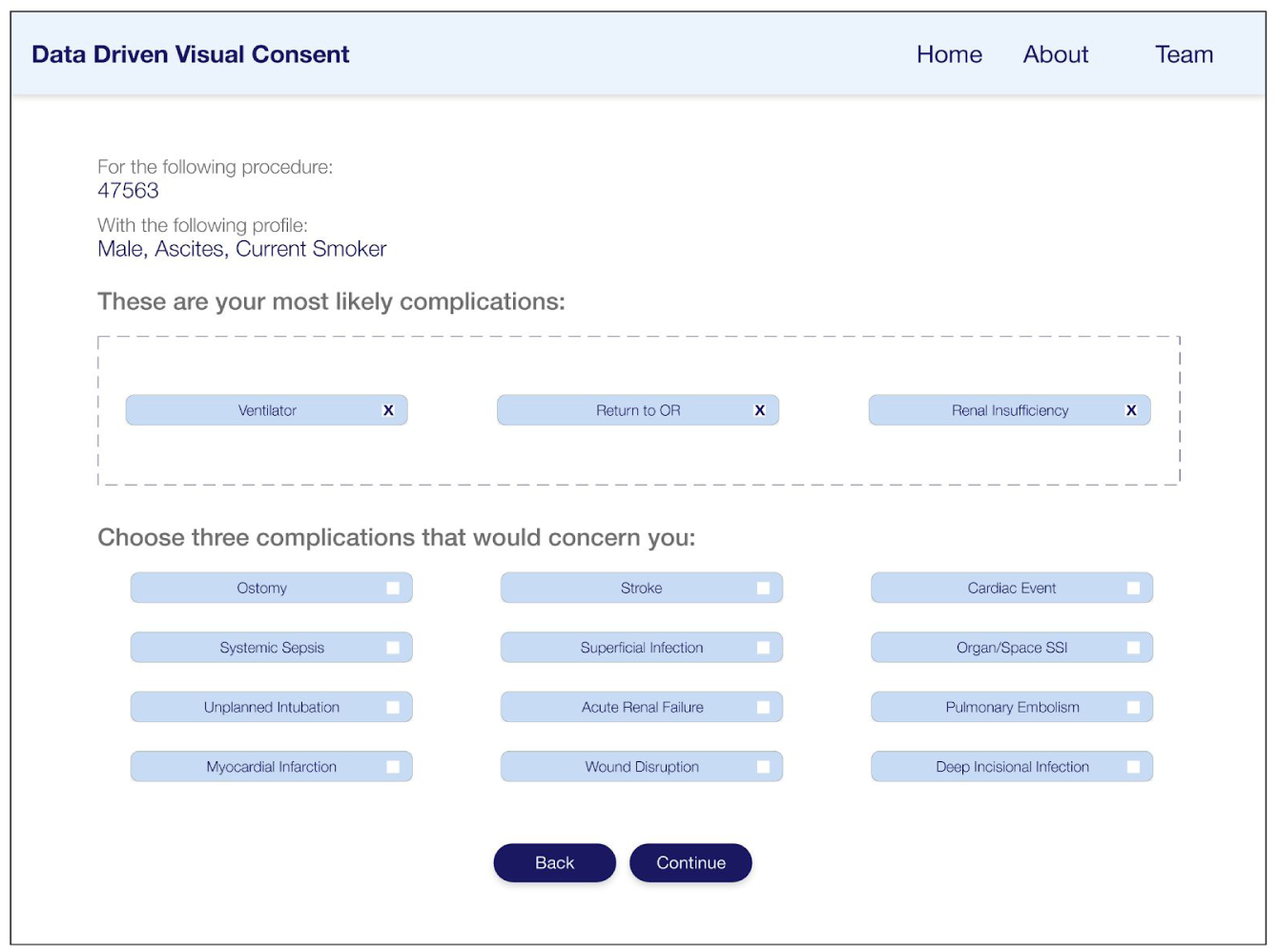
Incorporating Patient Preference. Three most common complications are pre-selected with the remaining complications listed in descending order of likelihood. The patient can select up to three risks at a time.

#### 3. Risk Visualization

The visualization is intended to communicate personalized perioperative risks and the likelihood of the discharge destinations in a clear and understandable manner. The overall goal is to promote a more coherent discussion between the surgeon and patient for improved shared decision making. The layout includes the most likely, pre-selected, complications based on the risk calculations as well as those selected by the patient and the likelihood of each discharge destination (Figure 4, top left). Discharge destinations are communicated using weighted lines to represent likelihood. Pre-selected and patient-selected complications are boxed separately to allow comparison between the two categories. Given the relatively low rates of complications, representation of the likelihood of each complication presented a unique challenge that we examined in detail.

**Figure 4:**
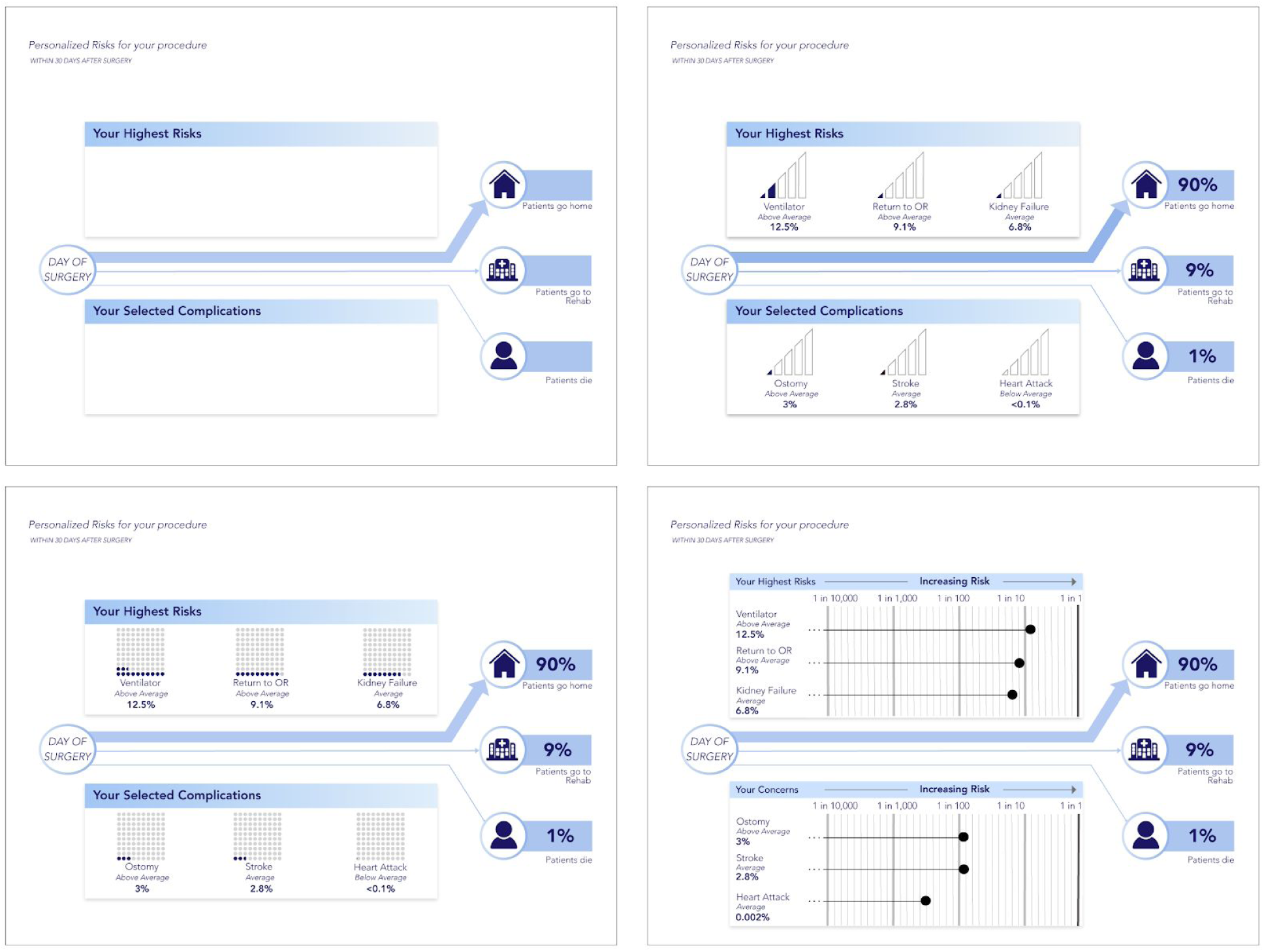
The design of the three risk visualizations used in this study. This study investigated three visualizations that, through a literature review, were identified as likely to be successful in communicating risks to patients. The top left shows the general layout that all three visualizations follow, where the patient’s highest risks are displayed on the top while the complications they have chosen are displayed below to allow for comparison. The likelihood of each discharge destination is separated from the risks and communicated using positional cues and weighted lines. The three visualizations tested were bar strength (top right), dot array (bottom left), and logarithmic scale visualization (bottom right).

We group complications into “rare events” (< 1%) and “common events” (>= 1%). We referred to the Visualizing Health repository (“Visualizing Health” n.d.) to choose visualizations that could be suitable for communicating these events. We chose the “bar strength” visualization that resembles the signal strength on mobile devices and represents a familiar visualization due to prevalence of mobile devices (Figure 4, top right). We also chose a waffle chart, called “dot array” because it is more granular than the bar strength and recommended by the Visualizing Health repository to accurately communicate risk (Figure 4, bottom left). To be able to show risks smaller than 1%, we chose a logarithmic scale inspired by the perspective scale proposed by Paling, which also allows for direct comparison of risks (Paling 2003) (Figure 4, bottom right).

### Evaluation of VCT

#### Participants

We conducted semi-structured interviews to evaluate these visualizations within the context of the VCT. A convenience sample of 20 patients was interviewed during their post-operative check up visit in the acute care surgical clinic at an academic hospital. The patients were approached by the interviewer and asked for their willingness to participate in the study. This study only included patients who had undergone a surgical intervention by an acute care surgeon and who agreed to participate with a written consent. The study was approved by the institutional review board (IRB) for human subjects research.

#### Interviewer

The Interviews were conducted by the first author of this paper. At the time of the study this researcher was holding a BS in Biomedical Engineering and completing their Master’s degree in Biomedical Informatics as a full time student and research assistant. This researcher didn’t have any prior relationship with the participants and only briefly stated the purpose of the study at the beginning of the interviews. The interviewer did not have any prior biases, aside from the assumption that visualization would be a useful tool for the consent process.

### Study Procedure

The interview guide was trialed with two individuals who were not participants in our study and refined to fit within 30 minutes as well as to cover all the essential topics. Figure 5 shows the structure of the final semi-structured interview (see Supplementary Information S1 for the interview guide). The interviews first aimed to understand the patient’s informed consent experience in the current practice - without any visualization aids (Figure 5, Part 1). The second section focused on risk perception and visualization preference (Figure 5, Part 2). Finally, the third section assessed perceptions of the value of a VCT and its usefulness during the informed consent process (Figure 5, Part 3). Each of the participants went through the interview only once and no repeat interviews were conducted.

**Figure 5:**
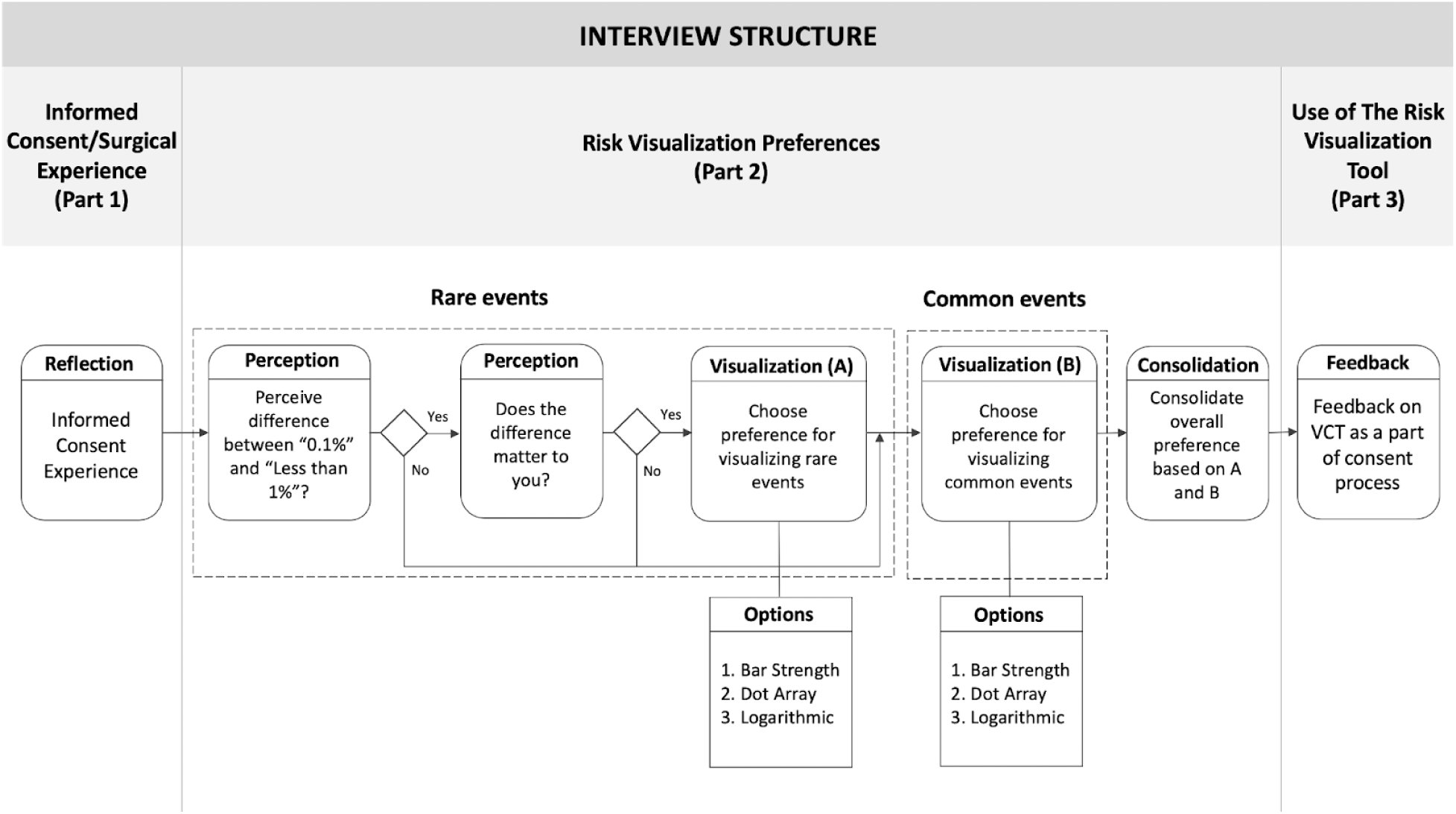
Semi-structured interview structure. Part 1 assessed the current informed consent experience. Part 2 assessed risk perception and risk visualization preference. Finally, Part 3 gathered feedback on the useful elements of VCT and its applicability.

The interviews were conducted in a clinical setting, after a postoperative visit. In some cases the interview was conducted in the presence of a significant other or a family member of the participant. The interviewer first gathered demographic data and impressions of the existing informed consent experience by recalling their most recent discussion about risks for informed consent with their surgeon. The interviewer then assessed the patient’s perception of the risk visualizations (Figure 5, Part 2). The interviewer evaluated perceptions of rare events by asking the patients if there was a notable difference between 0.1% and the phrase “less than 1%” when a likelihood of a rare event such as being struck by lightning was expressed. Patients who perceived a difference were shown three visualizations and asked to identify the visualization that conveyed the 0.1% chance of getting struck by lightning most clearly. The interviewer then assessed visualization preferences for common risks by using an example of a 12.3% chance of an earthquake. Again, the patient was shown the same three visualizations and asked to indicate which conveyed this information best. In cases where the visualizations chosen for rare events differed from the visualization chosen for common events, the patients choose one of the two preferred options for the final visualization. In the final step (Interview Structure, Part 3, see Figure 5), the full visualization was presented to the patient using the previously selected visualization type. To test the intuitiveness of the design, the patient was asked to explain what they saw and what decisions could be made without any explanation of what the visualization is trying to achieve. The interviewer then explained the intended purpose of the visualization and collected additional comments about the VCT and the patient was asked to identify situations in which they would find this tool useful. Additionally, the patient was asked what they found most useful, what could be improved, and any additional information they would like to see.

### Data Analysis

The interviews were recorded with an iPhone (Apple Inc, Cupertino, CA) and no field notes were taken during the interview. Interviews were transcribed using Dragon Dictate 3.0 (Nuance Corporation, Waltham, MA) with manual verification by the interviewer. The transcriptions were not sent back to the participants for comments and/or corrections. We conducted a mixed methods analysis of the data. For the qualitative part, the interviewer, together with another author of this paper, conducted thematic analysis of the data using Microsoft Word and Excel. Each of them independently inductively coded the transcribed interviews by assigning labels to the meaningful discourse units in the patients’ answers. After this, both researchers reviewed the codes and clarified the disagreements. Finally, in a collaborative manner, they grouped the codes into categories and the categories into themes. These were checked for validity with domain experts (two other authors of the paper) and carefully modified to accommodate their feedback. However, we did not conduct any member checks with the participants. We also applied a quantitative approach to the analysis and extracted discourse units that expressed different preferences and reasons for those preferences. We used these discourse units to assess the prevalence of different perceptions of the participants about the visualizations presented in our VCT.

## RESULTS

We interviewed 20 patients attending a postoperative visit. Average age of the cohort was 61.7 with a range of 29 to 87 (median = 59, sd =14) with 55% female and 45% male participants. The education level ranged from “some high school” to PhD. Most patients had gastrointestinal surgery of the intestines, gallbladder, or appendix, and about half of cases involved emergency procedures.

The thematic analysis of semi-structured interviews revealed three main categories: (1) *factors that influence risk perception*, (2) *perceptions of the visualizations*, and (3) *effects of the proposed VCT*.

### 1. Factors that Influence Risk Perception

We found that patients reacted positively to learning that the risks could be personalized. Some stated that the personalization of the risks was the highlight of the tool because it made them feel more carefully considered as a patient. We identified several factors that influence the perception of risk that encompass *the surgical procedure, the patient’s cognitive state* and *the timing of the consent*.

#### The Surgical Procedure

Factors that influence risk perception of the procedure included the clarity, complexity and urgency of the case. We found that most patients had a clear diagnosis and were confident with the surgeon’s familiarity with the case. They indicated that they were less concerned about risks associated with their surgery as compared to those patients with an unclear diagnosis. The latter made statements such as “[the surgeons] didn’t know what they were getting into” and some required multiple visits or surgeries. For patients who returned to the OR, all complications were of low importance and the pain or fear of death outweighed all others.

#### The Cognitive State of the Patient

The cognitive state proved to be an important factor for the perception of risk. Patients who were in pain or feeling drowsy cared less about complications and wanted to “get it over with.” A consistent theme was the delegation of decision-making to a more medically literate family member such as a spouse or child when such an opportunity existed. Patients with low health literacy were more likely to express uncertainty about diagnosis and felt that identification of complications is of low importance. For these patients, the opinion of the medical professional was of primary importance.

#### The Timing of Consent

The timing of consent varied between patients. For emergency cases, risks were communicated within a few hours of the surgery; for transfer cases or planned operations, risks could have been initially communicated a couple weeks in advance. For acute patients, lists of complications were of low importance because pain and death were described as important factors. Alternatively, sub-acute patients (surgeries within 24 hours but more than a few) felt that knowing the risks was important for expectation management. All respondents were clear that knowledge about risks would not have influenced their decision to go forward with surgery.

### 2. Factors that Influence Perceptions of the Visualizations

We assessed *how patients preferred rare events to be communicated, what visualization patients preferred*, and *whether comparison to the average was useful*. We also gathered feedback on the *overall impression of VCT*.

#### Preference for rare events communication

The majority of patients did not have a preference how rare events are communicated to them. Patients who wanted to know the exact percentage smaller than 1% (n=3) preferred the logarithmic representation for rare events.

#### Preference for Risk Visualization

Figure 6 is based on the preferred visualization after the participant was exposed to the visualization options for rare and common events and asked to consolidate their answer in a single visualization. Figure 6 shows preferences ranged across the visualizations. Of the three available graphics--the bar strength, dot array, and logarithmic scale--there was no consensus on a preferred visualization. Patients who preferred the bar strength visualization (n=5) liked the simplicity and clear step increases which allowed quick interpretation. Patients who liked this approach felt that it was less complicated than other options. One patient expressed concern over the discretization of the bars and a few patients felt it didn’t show enough information. The dot array was endorsed by four patients who preferred its visual organization and that it allowed comparison of ratios of shaded to grayed out dots. These patients found the dot array easy to understand and that it gave “just the right amount” of information. One patient noted that they would waste time counting dots and a few patients felt it would be hard to compare risks. Patients who preferred the logarithmic scale (n=7) felt that it communicated the risks most clearly, and allowed for easy comparison and aggregation of risks. Many mentioned that they liked the labelling of “1 in X”. However, patients who did not prefer the logarithmic scale found it the most complicated out of the three options.

**Figure 6:**
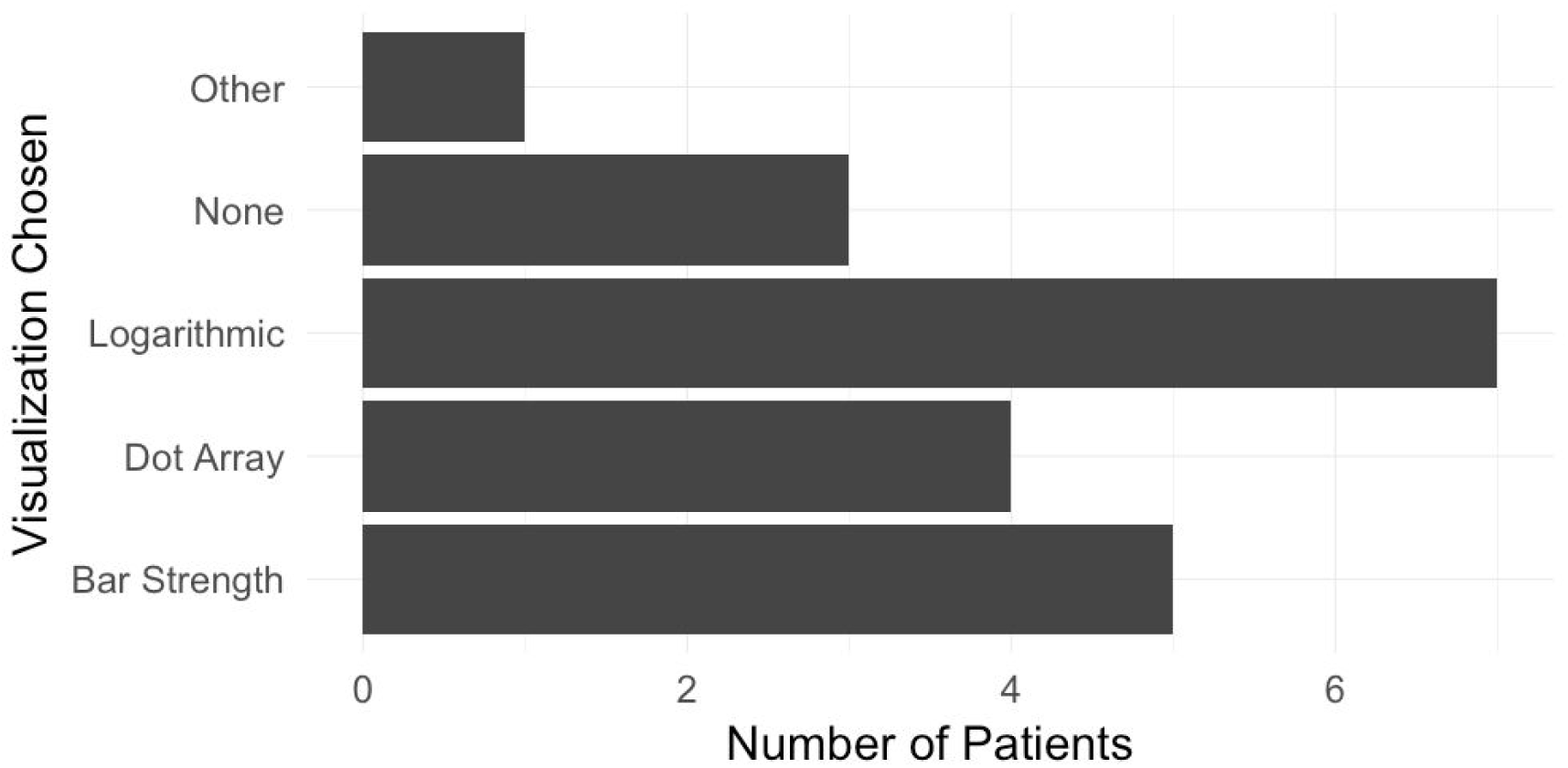
Visualization preferences of patients. The bar plot shows that patients had differing preferences for optimal visualization for communicating risks in the VCT. One participant liked a visual aid, but preferred a different visualization than the three presented. Three patients did not express interest in a visual aid.

Four patients did not respond as positively to the visualizations. One patient was unsatisfied with all of the choices presented. Three participants rejected visual aids altogether and preferred verbal communication or that the entire decision be left to the doctor. These patients were all over 75 years old which is notably higher than the average age of other participants.

#### Usefulness of comparison to the average

The majority of patients (61%) expressed indifference to knowing whether their risk was above or below average. For those that cared about the average, they stated that it would raise or lower their concern and some only cared if it was actionable information. Many were not confident in how to include this information in their decision making process.

#### Overall impression of VCT

In terms of intuitiveness, approximately half of the patients found the final visualization intuitive without any context. However, after explaining the context of the tool and the steps leading up to the final visualization, most patients felt able to make decisions based on the visualization.

### 3. Effects of VCT

We observed four major effects of the VCT on perceived informed consent: the *depth and length of the discussion, information retention*, and *risk awareness*.

#### Depth and Length of the Discussion

All of the patients agreed that VCT would help stimulate a deeper discussion with their provider. They claimed it would “help [them] think of new questions [they] hadn’t thought of before.’’ Most patients stated that it would have helped them pick up information more quickly and be more confident in their surgical decision. Patient’s claimed that VCT would allow them to have a better understanding of the possible complications and their likelihood. Some patients expressed concern that having this information and new questions may extend the discussion and would take too much of the surgeon’s time.

#### Information Retention

Most patients also felt that the visualizations may serve as a reference to consult after consent.

#### Risk Awareness

The majority of patients believed such a tool would make them more aware of potential risks. It made them more confident in their decision to pursue surgery and many noted that it would not have changed their decision to pursue surgery. A few patients noted that it prompted more long term thinking about what to expect after the surgery and how it would impact not only them but their families. Additionally, patients expressed concern that this may be too much information for other patients and it may dissuade other patients from pursuing a surgery that is in their best interest.

Patients expressed interest in seeing information generally not available in current risk calculators: pain level, length of hospital stay, and expected recovery time. Our interviews revealed that patients were most concerned about their potential health status and whether they would be able to continue normal activity after surgery--including the chances of avoiding an ostomy (Figure 7).

**Figure 7:**
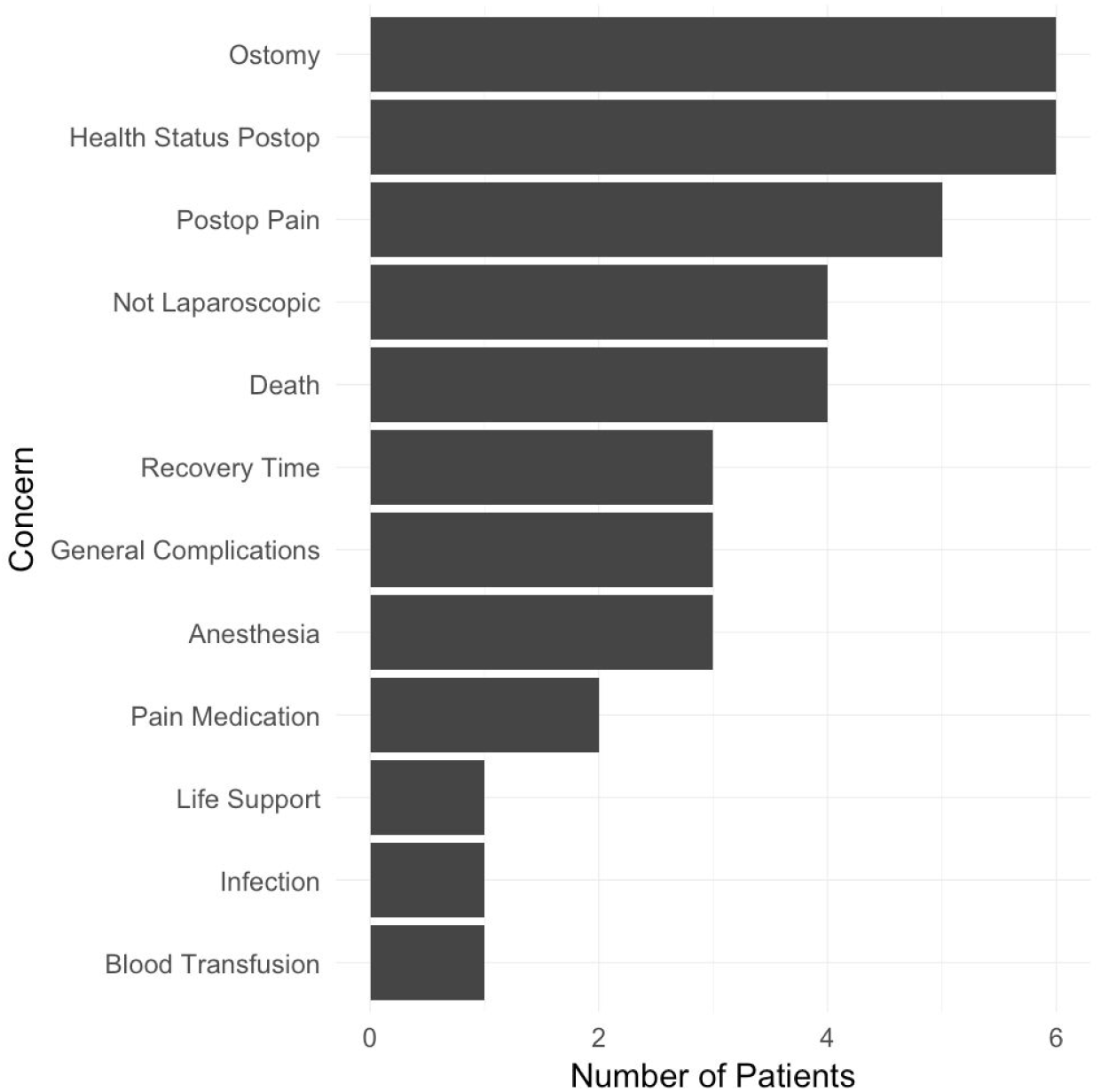
The major concerns of patients before surgery. This bar plot reveals that patients were most concerned about their potential health status, possible ostomy, and the pain level they could expect after surgery.

## DISCUSSION

Our study is the first to enumerate the opportunities and requirements of a personalized visual consent form for surgery. Most importantly, we found that patients’ main concerns did not align with the highest risks or, in some cases, what traditional risk calculators consider to be a risk worth communicating. Second, we found that a visual aid was perceived by patients to improve information retention and risk awareness. However, our interviews could not identify a universal visualization method; patients have different preferences for the type of risk visualization. Overall, we believe that a Visual Consent Tool (VCT) can improve shared decision making and be beneficial for patients and providers, if appropriately customized to the particular context pertinent to the patient.

While personalization was appreciated by participants, we uncovered multiple limitations to current surgical risk calculators. We found that patient’s main concerns did not always align with the patient’s highest risks or with the content of the most common calculator. Future work should consider expanding surgical risk calculations to include patient concerns such as pain levels, family burden, or functional outcomes. Additionally, a limitation of all current surgical risk calculators is their inability to calculate and communicate risks associated with not pursuing a surgical intervention.

Almost all patients agreed that the visualizations were helpful in understanding risks. However, there was not a single visualization that was preferable for the majority of patients. With our limited dataset, these preferences could not be reliably mapped to demographic indicators such as age or education level, for example. This aligns with the findings of previous studies, which indicated that communicating risk in a tailored way leads to higher satisfaction (Bickmore et al. 2015). In this case, we were not able to identify a universal visualization approach. Furthermore, we did not assess the level of understanding and recall for the different visualizations. Since preference and understanding may not align, future studies should include quantitative measures of retention by preferred visualization. These measures should include the factors that influence risk perception described in our study.

Our findings should be considered in light of the limitations of this study. The study population is biased towards older patients and only includes patients who underwent a specific subgroup of general surgeries. Their relatively positive experiences and their historical exposure may have influenced their recall and opinions. Patients who undergo different surgeries, have a different demographic makeup, have worse outcomes, or are in the pre-operative period may have very different risk perceptions and risk visualization preferences than our study population.

Another important consideration are the stakeholders involved in incorporating VCT into the current clinical workflow. These stakeholders include the patient and their family, surgeons, and hospital administration. This study focuses on the preference of the patients but future work should consider input from surgeons and hospital administrators to find a solution that maximizes benefits for all. For example, while exhibiting benefits for the patients, some patients expressed concern that the interactions stimulated by the introduction of the VCT might take too much of the surgeon’s time and negatively affect their clinical productivity. Future work should consider how to enable VCT-based communication efficiency that will benefit both patients and surgeons, and not significantly favor one over the other. While the VCT may disrupt current practices, it is also important to consider the greater value of such a tool to the hospital and administration. Most patients agreed that the VCT raises awareness, stimulates new questions, and allows them to reflect on the discussion with their surgeon after the conversation. Participants believed these benefits would allow them to take a more active role in their treatment plan. For these reasons, we anticipate that the proposed method will help promote shared decision making by decreasing the opportunities for miscommunication and making patients more confident in their decisions. When patients are more comprehensively informed they are less likely to pursue legal action when they experience a non-beneficial outcome (Grauberger et al. 2017). For these reasons we believe that VCT not only shows promise for patients but for the healthcare system as a whole.

In conclusion, we found that there is no universal way of visually communicating risks to patients, which counters the current practice of using a one size fits all approach. We found that multiple factors impact the perception of risks and that the proposed VCT has potential to provide useful information to patients and to stimulate shared decision making with their surgeons. We anticipate that these benefits can be achieved if patient characteristics are taken into account to deliver a tailored risk visualization solution. Finally, we postulate that the need for tailored visual communication of complex medical information applies to other domains of healthcare as well.

## Data Availability

No sharable data was collected.

## ACKNOWLEDGEMENTS

We thank Maura Cass from IDEO for valuable feedback on our evaluation study as well as Drs. Parsons, Cook, and Hauser at BIDMC for providing access to their patients. Finally, we acknowledge Shannon Ehmsen at Harvard Medical School for help with graphic design of the visualizations.

## SUPPLEMENTARY INFORMATION

S1. Semi-structured Interview Guide

## REFERENCES

Angelos, Peter. 2012. “The Evolution of Informed Consent for Surgery Using the Best Case/Worst Case Framework.” Internal Medicine 27 (10): 1361–67.

Bertsimas, Dimitris, Jack Dunn, George C. Velmahos, and Haytham M. A. Kaafarani. 2018. “Surgical Risk Is Not Linear: Derivation and Validation of a Novel, User-Friendly, and Machine-Learning-Based Predictive OpTimal Trees in Emergency Surgery Risk (POTTER) Calculator.” Annals of Surgery 268 (4): 574–83.

Bickmore, Timothy, Dina Utami, Shuo Zhou, Candace Sidner, Lisa Quintiliani, and Michael K. Paasche-Orlow. 2015. “Automated Explanation of Research Informed Consent by Virtual Agents.” In Lecture Notes in Computer Science, 260–69.

Bilimoria, Karl Y., Yaoming Liu, Jennifer L. Paruch, Lynn Zhou, Thomas E. Kmiecik, Clifford Y. Ko, and Mark E. Cohen. 2013. “Development and Evaluation of the Universal ACS NSQIP Surgical Risk Calculator: A Decision Aid and Informed Consent Tool for Patients and Surgeons.” Journal of the American College of Surgeons 217 (5): 833–42.e1–3.

Brennan, Meghan, Sahil Puri, Tezcan Ozrazgat-Baslanti, Zheng Feng, Matthew Ruppert, Haleh Hashemighouchani, Petar Momcilovic, Xiaolin Li, Daisy Zhe Wang, and Azra Bihorac. 2019. “Comparing Clinical Judgment with the MySurgeryRisk Algorithm for Preoperative Risk Assessment: A Pilot Usability Study.” Surgery, February. https://doi.org/10.1016/j.surg.2019.01.002.

Cooper, Zara, Andrew Courtwright, Ami Karlage, Atul Gawande, and Susan Block. 2014. “Pitfalls in Communication That Lead to Nonbeneficial Emergency Surgery in Elderly Patients With Serious Illness: Description of the Problem and Elements of a Solution.” Annals of Surgery 260 (6): 949–57.

Fink, Aaron S., Allan V. Prochazka, William G. Henderson, Debra Bartenfeld, Carsie Nyirenda, Alexandra Webb, David H. Berger, et al. 2010. “Predictors of Comprehension during Surgical Informed Consent.” Journal of the American College of Surgeons 210 (6): 919–26.

Grauberger, Jennifer, Panagiotis Kerezoudis, Asad J. Choudhry, Mohammed Ali Alvi, Ahmad Nassr, Bradford Currier, and Mohamad Bydon. 2017. “Allegations of Failure to Obtain Informed Consent in Spinal Surgery Medical Malpractice Claims.” JAMA Surgery 152 (6): e170544.

Ingraham, Angela M., Suresh K. Agarwal, Hee Soo Jung, Amy E. Liepert, Ann P. O’Rourke, and John E. Scarborough. 2017. “Patient-Centered Outcome Spectrum: An Evidence-Based Framework to Aid in Shared Decision-Making.” Annals of Surgery, September. https://doi.org/10.1097/SLA.0000000000002466.

Kruser, Jacqueline M., Michael J. Nabozny, Nicole M. Steffens, Karen J. Brasel, Toby C. Campbell, Martha E. Gaines, and Margaret L. Schwarze. 2015. “‘Best Case/Worst Case’: Qualitative Evaluation of a Novel Communication Tool for Difficult in-the-Moment Surgical Decisions.” Journal of the American Geriatrics Society 63 (9): 1805–11.

Meguid, Robert A., Michael R. Bronsert, Elizabeth Juarez-Colunga, Karl E. Hammermeister, and William G. Henderson. 2016. “Surgical Risk Preoperative Assessment System (SURPAS): III. Accurate Preoperative Prediction of 8 Adverse Outcomes Using 8 Predictor Variables.” Annals of Surgery 264 (1): 23–31.

“NQF: Surgery 2015-2017 Final Report.” n.d. Accessed August 21, 2019. https://www.qualityforum.org/Publications/2017/04/Surgery_2015-2017_Final_Report.aspx.

Paling, John. 2003. “Strategies to Help Patients Understand Risks.” BMJ 327 (7417): 745–48.

Scheer, Adena S., Annette M. O’Connor, Beverly P. K. Chan, Husein Moloo, Eric C. Poulin, Joseph Mamazza, Rebecca C. Auer, and Robin P. Boushey. 2012. “The Myth of Informed Consent in Rectal Cancer Surgery: What Do Patients Retain?” Diseases of the Colon and Rectum 55 (9): 970–75.

Taylor, Lauren J., Michael J. Nabozny, Nicole M. Steffens, Jennifer L. Tucholka, Karen J. Brasel, Sara K. Johnson, Amy Zelenski, et al. 2017. “A Framework to Improve Surgeon Communication in High-Stakes Surgical Decisions: Best Case/Worst Case.” JAMA Surgery 152 (6): 531–38.

Tong, Allison, Peter Sainsbury, and Jonathan Craig. 2007. “Consolidated Criteria for Reporting Qualitative Research (COREQ): A 32-Item Checklist for Interviews and Focus Groups.” International Journal for Quality in Health Care: Journal of the International Society for Quality in Health Care / ISQua 19 (6): 349–57.

“Visualizing Health.” n.d. Accessed December 16, 2019. http://www.vizhealth.org/.

